# Switching Between VCTE and 2D-SWE Reclassifies Fibrosis Risk in Metabolic Dysfunction-Associated Steatotic Liver Disease

**DOI:** 10.64898/2026.07.28.26359104

**Authors:** Vasily Isakov, Alexei Goncharov, Murad Israpilov

**Affiliations:** Department of Gastroenterology, Hepatology & Nutrition, Biotechnology & Food Safety, Moscow, Russia; Department of Radiology of Federal Research Center of Nutrition, Biotechnology & Food Safety, Moscow, Russia

**Author notes:** **Correspondence:** Vasily Isakov, MD, PhD Department of Gastroenterology, Hepatology & Nutrition, Moscow, Russia.

**Keywords:** metabolic dysfunction-associated steatotic liver disease, elastography, method comparison, agreement

## Abstract

**Background and Aims:** Vibration-controlled transient elastography (VCTE) and two-dimensional shear-wave elastography (2D-SWE) are used to assess liver stiffness in metabolic dysfunction-associated steatotic liver disease (MASLD); patients may be assessed by different methods over time. Because 2D-SWE fibrosis cut-offs are not uniformly validated and no biopsy or magnetic resonance elastography reference is available, we asked whether switching methods moves patients across decision thresholds. We evaluated agreement and decision-threshold interchangeability between VCTE and 2D-SWE in MASLD with obesity.

**Methods:** In a retrospective cross-sectional agreement study at a tertiary-care center, 317 consecutive adults with MASLD underwent same-day VCTE (FibroScan) and GE LOGIQ E9/E10 2D-SWE. Continuous agreement was assessed by Bland–Altman analysis; the categorical analysis compared binary clinical decision thresholds (VCTE ≥8.0/≥10.0 kPa; 2D-SWE ≥7.204/≥8.060 kPa) using Cohen κ and McNemar tests. Prespecified sensitivity analyses and a post-hoc recalibration were performed; VCTE served as operational reference.

**Results:** VCTE yielded higher values (geometric mean VCTE/2D-SWE ratio, 1.23; ratio limits of agreement, 0.64–2.40; intraclass correlation coefficient, 0.41). At the lower threshold, 71 of 117 VCTE-positive patients (61%) were below the 2D-SWE threshold versus 5 of 200 (2.5%) reclassified upward (agreement 76.0%; κ 0.417; P<.001); the upper threshold was similar (38 of 63, 60%, vs 8 of 254, 3.1%). Discordance persisted across cut-offs; agreement worsened descriptively across BMI strata. Recalibration removed the directional asymmetry but not the discordance (agreement unchanged, 76.0%).

**Conclusions:** In MASLD patients, switching from VCTE to 2D-SWE reclassified decision-threshold status in ∼60% of VCTE-positive patients; recalibration removed the directional bias but not the disagreement. Follow-up should use the same elastography modality and, when possible, the same platform.

## Introduction

Metabolic dysfunction-associated steatotic liver disease (MASLD) affects approximately 30% of adults worldwide.^1, 2^ The risks of all-cause and liver-related mortality increase with fibrosis stage; ^3, 4^ therefore, timely fibrosis assessment is a cornerstone of risk stratification and treatment decision-making. Because liver biopsy is unsuitable for repeated population-level assessment, contemporary clinical pathways increasingly rely on noninvasive tests to identify patients at risk of advanced fibrosis and to support serial reassessment. Imaging-based liver stiffness measurement has consequently become an important component of scalable fibrosis risk stratification in patients with MASLD, including the growing population with severe obesity.^5, 6^

Vibration-controlled transient elastography (VCTE) and two-dimensional shear-wave elastography (2D-SWE) are established ultrasound-based methods for measuring liver stiffness. Although both report results in kilopascals, they differ in wave generation, image guidance, sampling geometry, acquisition technique, and manufacturer-specific calibration.^7^ Diagnostic performance or correlation between methods therefore does not necessarily establish numerical interchangeability at the individual-patient level. This distinction is clinically important because patients may undergo follow-up using different techniques or devices as they move between hepatology, radiology, metabolic, and bariatric services.

Comparative studies across several 2D-SWE platforms have reported strong correlations with VCTE, similar diagnostic performance against histology, or good reproduction of VCTE-defined fibrosis-risk categories.^8–13^ However, these findings do not establish numerical interchangeability: most studies prioritized diagnostic discrimination or categorical concordance, and studies reporting Bland–Altman analyses found wide limits of agreement. Directional reclassification at MASLD-relevant thresholds in a predominantly severely obese cohort has not been specifically evaluated.

This evidence gap has practical consequences. Switching elastography methods during longitudinal follow-up may produce an apparent change in liver stiffness that reflects the measurement platform rather than disease progression or regression. A patient may also cross a threshold used to prompt additional evaluation or closer follow-up solely because a different technique was used. This is especially relevant in bariatric and metabolic practice, where severe obesity can affect measurement performance and where patients commonly move between services and imaging platforms. Agreement should therefore be evaluated using patient-level limits of agreement and directional threshold reclassification, rather than inferred from correlation, similar group-level distributions, or area under the receiver-operating-characteristic curve alone.

We therefore evaluated agreement between same-day VCTE and 2D-SWE liver stiffness measurements in adults with MASLD. The objectives were to quantify continuous patient-level agreement and proportional bias; determine concordance and directional reclassification at clinically relevant VCTE-referenced decision thresholds; and explore clinical and technical predictors of absolute and signed discrepancies between methods. We additionally examined the robustness of agreement across alternative thresholds, reliability criteria, VCTE probe type, 2D-SWE device model, and body mass index strata. VCTE was used as the operational reference for these comparisons, rather than as a diagnostic gold standard.

## Methods

### Study Design and Participants

This single-center, retrospective, cross-sectional agreement study included consecutive adults with MASLD (89% with obesity) who underwent same-day VCTE and 2D-SWE during routine assessment at the Department of Gastroenterology, Hepatology and Nutrition, Federal Research Center of Nutrition, Biotechnology and Food Safety, Moscow, Russia. MASLD was defined by hepatic steatosis on abdominal ultrasound, alcohol consumption below 20 g/day in women and 30 g/day in men, negative HBsAg and anti-HCV antibodies, no clinical, biochemical, or serological evidence of another chronic liver disease, and at least one cardiometabolic risk factor according to contemporary criteria.^14^ Eligible patients had valid paired LSMs obtained under standardized fasting conditions. Of 335 paired examination records assessed, 17 failed prespecified reliability criteria (16 VCTE and 1 2D-SWE), leaving 318 records. One patient underwent 2D-SWE twice on the same day, once on each LOGIQ platform. To retain one observation per patient, the first examination (LOGIQ E9) was used in the primary analysis (n=317); both examinations were retained only in the prespecified device-model sensitivity analysis (n=318).

The study followed the Guidelines for Reporting Reliability and Agreement Studies (GRRAS) and a written statistical analysis plan (SAP). This was a retrospective observational method-comparison study in which participants were not prospectively assigned to an intervention or comparison group; therefore, it did not meet the ICMJE definition of a clinical trial, and clinical-trial registration and CONSORT reporting were not applicable. The two post-hoc analyses (the Deming-regression leverage analysis and operational recalibration analysis) were date-stamped after completion of the primary analyses and documented in a separate amendment. The SAP and amendment were date-stamped, archived on the Open Science Framework, and provided as Supplementary Material.

The study was conducted in accordance with the Declaration of Helsinki and was approved by the Ethics Committee of the Federal Research Center of Nutrition, Biotechnology and Food Safety, Moscow, Russia (protocol No. 4; April 10, 2026). The committee waived the requirement for separate study-specific informed consent because the study involved retrospective analysis of data obtained during routine clinical care, included no additional interventions or study-specific procedures, and used an anonymized analytical database.

### Elastography Measurements

VCTE was performed using the FibroScan 530 (Echosens, Paris, France), with the M or XL probe selected by the automated probe-selection tool and simultaneous measurement of the controlled attenuation parameter. Reliability was classified according to the Boursier criteria: examinations were considered poorly reliable when the interquartile range/median was >30% and median LSM was ≥7.1 kPa; examinations with an interquartile range/median >30% but median LSM <7.1 kPa were retained.^15^ Poorly reliable examinations were excluded before database lock. 2D-SWE was performed on GE LOGIQ E9 or E10 systems (GE HealthCare, USA). Because validated LOGIQ-specific reliability rules are not established for 2D-SWE, an IQR/median ≤30% criterion was used as the prespecified reliability criterion. Liver stiffness was expressed in kilopascals for both modalities.

Both examinations were performed after at least 8 hours of fasting by separate experienced operators who were unaware of the other modality’s result. VCTE was performed according to the manufacturer’s protocol, with at least 10 valid acquisitions and the median reported as the final LSM. For 2D-SWE, measurements were obtained through a right intercostal approach in the right hepatic lobe during a brief breath hold. The acquisition region of interest was positioned at least 1 cm below the liver capsule and no more than 6 cm from the skin surface in homogeneous parenchyma free of large vessels. An individual acquisition was considered invalid when <50% of the acquisition region was filled by the color-coded elastography map.^16, 17^ Twelve valid acquisitions were obtained, and their median was used as the final 2D-SWE LSM. Age, sex, body mass index (BMI), waist circumference, controlled attenuation parameter, alanine aminotransferase, and measurement-quality indices were recorded at the same visit.

### Clinical Decision Thresholds

Because no biopsy or magnetic resonance elastography reference was available and complete histologic staging thresholds are not uniformly validated across 2D-SWE platforms, categorical agreement was evaluated at binary clinical decision thresholds rather than by assigning F0–F4 stages. The prespecified VCTE thresholds were ≥8.0 and ≥10.0 kPa, rounded from the commonly used 7.9-kPa rule-out and 9.6-kPa rule-in thresholds for advanced fibrosis.^7, 18^ These values are also clinically relevant starting points for longitudinal VCTE risk assessment.^19, 20^ The paired 2D-SWE thresholds (7.204 and 8.060 kPa) were the most applicable published GE-specific MASLD estimates from the recent MASLD meta-analysis.^21^ The exact 9.6-kPa VCTE threshold and alternative published frameworks were examined in sensitivity analyses. VCTE served as an operational reference for categorical comparisons and was not considered diagnostic truth.

### Statistical Analysis

Continuous variables were summarized as median [interquartile range] and categorical variables as number (percentage). The co-primary analyses were Bland–Altman assessments of VCTE minus 2D-SWE on the original kPa scale and, because heteroscedasticity was expected, on the natural-log scale. The latter was back-transformed to obtain the geometric mean VCTE/2D-SWE ratio and ratio limits of agreement. Bias and 95% limits of agreement were reported with 95% confidence intervals. Proportional bias was examined by regressing the intermethod difference on the paired mean and interpreted descriptively because of mathematical coupling between the difference and mean. Additional continuous-agreement analyses included the intraclass correlation coefficient (two-way random effects, absolute agreement, single measures), Lin concordance correlation coefficient, Deming regression, and Passing–Bablok regression.

Categorical agreement was evaluated using 2×2 tables, overall, positive, and negative agreement, Cohen κ, and the McNemar test of discordance asymmetry. Bias-corrected and accelerated bootstrap confidence intervals were calculated for κ in the primary analyses. Discordant pairs were reported separately as VCTE-positive/2D-SWE-negative and VCTE-negative/2D-SWE-positive. Robustness was examined across alternative published thresholds, including all-vendor MASLD,^21^ GE LOGIQ technical white-paper thresholds,^22^ and LOGIQ E9 NAFLD/high-BMI cut-offs,^17^ borderline-zone exclusions, stricter reliability requirements, VCTE probe, 2D-SWE platform, and BMI strata; full specifications are provided in the SAP and Supplementary Tables. Bootstrap confidence intervals for Bland–Altman bias and limits of agreement were used for small strata (n <60) or when the distribution of within-stratum differences was questionable.

Exploratory correlates of absolute and signed intermethod discrepancy were assessed using ordinary least-squares regression with heteroscedasticity-consistent standard errors and restricted cubic splines. Candidate variables included age, sex, BMI, waist circumference, controlled attenuation parameter, alanine aminotransferase, probe, platform, measurement-quality indices, and mean LSM. Benjamini–Hochberg false-discovery-rate-adjusted q values were reported for exploratory testing.

Two analyses were conducted post hoc. First, the leverage sensitivity of the Deming slope was examined using exclusion, winsorization, restricted-range, influence, jackknife, and error-variance-ratio scenarios, with Passing–Bablok regression as an outlier-robust comparator. Second, an operational recalibration analysis derived LOGIQ 2D-SWE thresholds that best reproduced the VCTE decision-threshold labels using receiver-operating-characteristic analysis, with the area under the curve estimated using DeLong confidence intervals and Youden-based cut-offs as the primary recalibrated thresholds; Passing–Bablok and Deming regression-based threshold inversion were used as cross-checks. Internal validation used optimism-corrected bootstrapping, and a paired bootstrap re-derived the cut-off within each resample to estimate confidence intervals for changes in agreement, κ, the number of VCTE-positive/2D-SWE-negative discordant pairs, and directional discordance proportion. The recalibrated thresholds were interpreted as candidate operational harmonization thresholds and not as diagnostic cut-offs for histological fibrosis.

All tests were two-sided, with emphasis on effect sizes and 95% confidence intervals. Analyses were performed using R version 4.5.2 (R Foundation for Statistical Computing, Vienna, Austria).

## Results

### Study Population

After measurement-quality exclusions and removal of one duplicated patient record, 317 unique patients were included in the primary analysis; the device-model sensitivity analysis included 318 examinations because both same-day 2D-SWE examinations from the duplicated case were retained. All analysis variables were complete, and no imputation was required (Supplementary Table S1). The cohort included 229 women (72%), with median age 57 years [interquartile range, 49–63] and BMI 37 kg/m² [33–43]. Overall, 281 patients (88.6%) had BMI ≥30 kg/m² and 195 (61.5%) had BMI ≥35 kg/m². The XL probe was used in 237 patients (75%) and the LOGIQ E9 platform in 216 (68%). Median LSM was 6.3 kPa [5.0–9.1] by VCTE and 5.45 kPa [4.61–6.35] by 2D-SWE. Patients with VCTE ≥8.0 kPa had higher BMI, waist circumference, and controlled attenuation parameter and were more frequently examined with the XL probe and LOGIQ E9 platform (Table 1).

**Table 1.** Baseline Characteristics of the Study Population.

| Characteristic | Overall (N = 317) | VCTE < 8.0 kPa (n = 200) | VCTE ≥ 8.0 kPa (n = 117) | P value |
| --- | --- | --- | --- | --- |
| Age, y | 57 (49–63) | 56 (49–62) | 58 (50–64) | .092 |
| Sex |  |  |  | .20 |
| Male | 88 (27.8%) | 61 (30.5%) | 27 (23.1%) |  |
| Female | 229 (72.2%) | 139 (69.5%) | 90 (76.9%) |  |
| Body mass index, kg/m<sup>2</sup> | 37 (33–43) | 35 (32–40) | 41 (36–46) | < .001 |
| Waist circumference, cm | 107 (98–118) | 103 (97–112) | 114 (105–124) | < .001 |
| CAP, dB/m | 307 (288–330) | 301 (281–326) | 318 (302–338) | < .001 |
| ALT, U/L | 26 (19–40) | 26 (19–36) | 27 (20–44) | .20 |
| VCTE LSM, kPa | 6.3 (5.0–9.1) | 5.2 (4.7–6.2) | 10.4 (8.8–12.3) | < .001 |
| 2D-SWE LSM, kPa | 5.45 (4.61–6.35) | 5.04 (4.38–5.77) | 6.59 (5.56–8.16) | < .001 |
| VCTE probe type |  |  |  | .044 |
| M | 80 (25.2%) | 58 (29.0%) | 22 (18.8%) |  |
| XL | 237 (74.8%) | 142 (71.0%) | 95 (81.2%) |  |
| LOGIQ model |  |  |  | < .001 |
| E9 | 216 (68.1%) | 118 (59.0%) | 98 (83.8%) |  |
| E10 | 101 (31.9%) | 82 (41.0%) | 19 (16.2%) |  |
| VCTE IQR/median, % | 22 (15–28) | 20 (14–27) | 24 (18–28) | .072 |
| 2D-SWE IQR/median, % | 15 (11–20) | 15 (10–19) | 16 (11–21) | .20 |
| VCTE Boursier reliability class |  |  |  | < .001 |
| Very reliable | 27 (8.5%) | 25 (12.5%) | 2 (1.7%) |  |
| Reliable | 290 (91.5%) | 175 (87.5%) | 115 (98.3%) |  |
| Poorly reliable | 0 (0%) | 0 (0%) | 0 (0%) |  |

Data are median (interquartile range) or n (%). P values compare patients with VCTE <8.0 versus ≥8.0 kPa using the Wilcoxon rank-sum or chi-squared test, as appropriate.

### Primary Continuous Agreement

VCTE exceeded 2D-SWE by a mean of 1.91 kPa (95% CI, 1.52–2.30), with wide 95% limits of agreement (−4.97 to 8.78 kPa). On the natural-log scale, the geometric mean VCTE/2D-SWE ratio was 1.234 (95% CI, 1.19–1.28), with ratio limits of agreement of 0.64–2.40. The difference increased with mean stiffness (slope, 0.998; 95% CI, 0.925–1.071; R²=0.70), although this association was interpreted descriptively because the difference and mean are mathematically coupled. Overall agreement was limited (ICC[2,1], 0.41; 95% CI, 0.21–0.56; Lin concordance correlation coefficient, 0.41; 95% CI, 0.36–0.46). Deming (slope, 3.40; 95% CI, 2.94–3.86) and Passing–Bablok (slope, 2.47; 95% CI, 2.15–2.87) regression were incompatible with the line of identity (Table 2; Figure 1).

**Figure 1.**
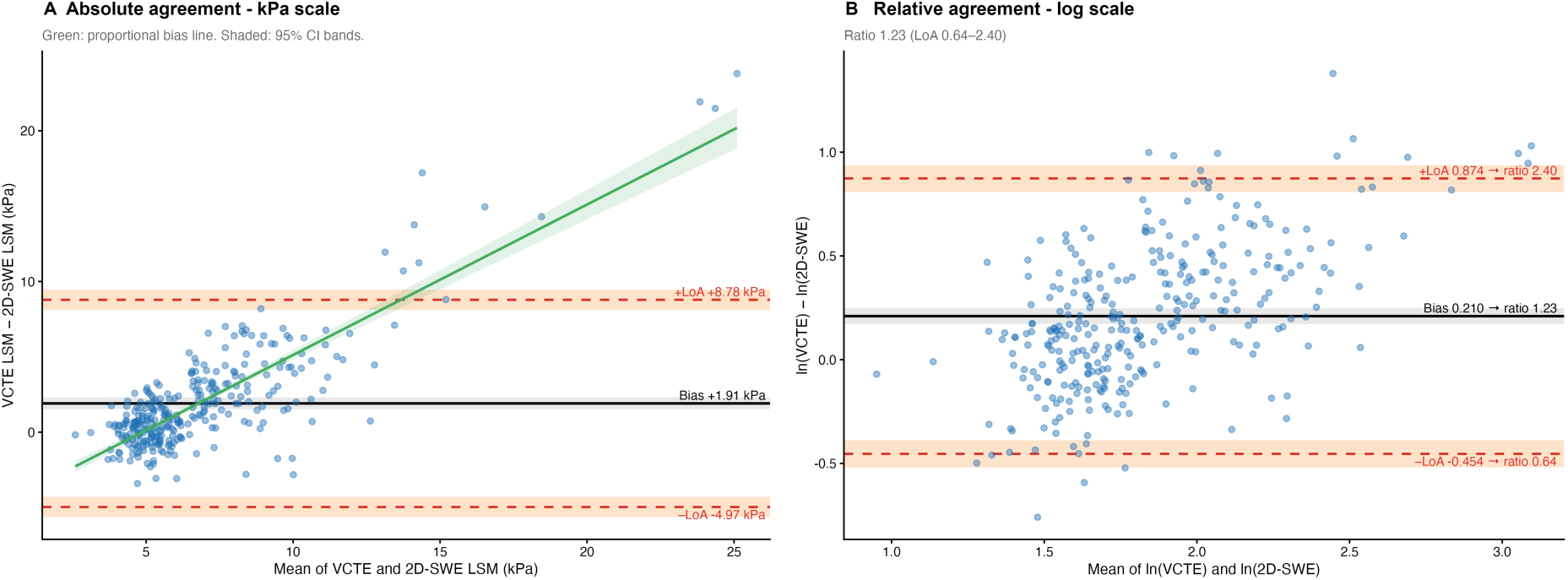
Bland–Altman agreement between paired VCTE and 2D-SWE liver-stiffness measurements on the absolute kPa (A) and log-ratio (B) scales. Solid black lines indicate mean bias, dashed red lines the 95% limits of agreement, and shaded bands their 95% confidence intervals. The green line indicates proportional bias.

**Table 2.** Continuous Agreement Between VCTE and 2D-SWE (N = 317)

| **Metric** | **Estimate** | **95% CI** |
| --- | --- | --- |
| Bias (VCTE – 2D-SWE), kPa | 1.91 | 1.52 to 2.30 |
| Lower limit of agreement, kPa | –4.97 | –5.64 to –4.30 |
| Upper limit of agreement, kPa | 8.78 | 8.11 to 9.46 |
| Geometric mean ratio (VCTE/2D-SWE) | 1.23 | 1.19 to 1.28 |
| Ratio limits of agreement | 0.64 to 2.40 | — |
| Intraclass correlation coefficient | .41 | .21 to .56 |
| Lin concordance correlation coefficient | .41 | .36 to .46 |
| Deming regression slope | 3.40 | 2.94 to 3.86 |
| Deming regression intercept | –12.05 | –14.56 to –9.55 |
| Passing–Bablok slope | 2.47 | 2.15 to 2.87 |
| Passing–Bablok intercept | –6.79 | –8.98 to –5.02 |

Bias is the mean of (VCTE – 2D-SWE). A regression slope different from 1 and an intercept different from 0 indicate that the two methods do not measure on the same scale. The intraclass correlation coefficient is ICC(2,1) (two-way random effects, absolute agreement, single measures).

### Agreement at Clinical Decision Thresholds

At the lower threshold, overall agreement was 76.0% (κ, 0.417; 95% CI, 0.32–0.51): 71 of 117 VCTE-positive patients (60.7%) were below the paired 2D-SWE threshold, compared with 5 patients classified in the reverse direction (McNemar P<.001). At the upper threshold, agreement was 85.5% (κ, 0.445; 95% CI, 0.31–0.57), with 38 of 63 VCTE-positive patients (60.3%) below the 2D-SWE threshold compared with 8 in the reverse direction (P<.001). The exact 9.6-kPa VCTE sensitivity analysis showed the same directional pattern (42 vs 7; P<.001) (Table 3; Figure 2).

**Figure 2.**
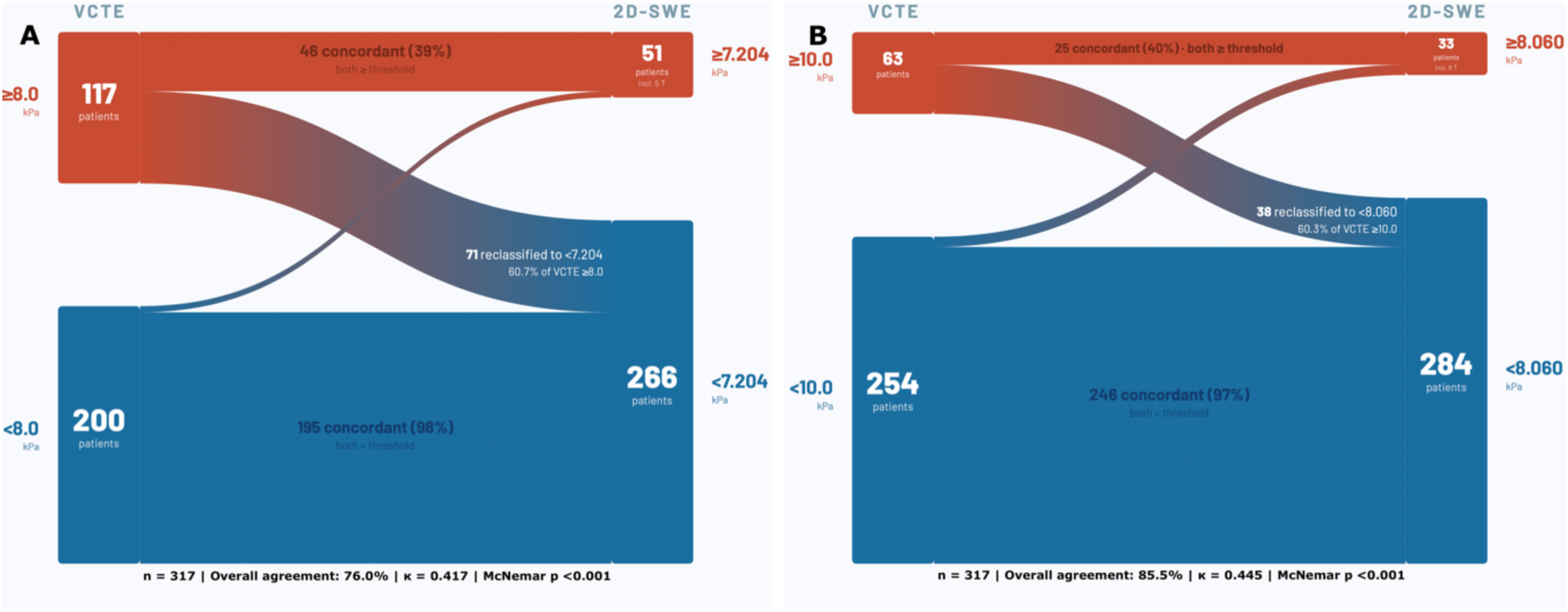
Agreement and directional reclassification at the lower (A) and upper (B) clinical decision thresholds. Crossing flows indicate discordant classifications. VCTE was used as an operational reference; categories represent device-specific decision-threshold classifications rather than histologic fibrosis stages.

**Table 3.** Agreement Between VCTE and 2D-SWE at the Clinical Decision Thresholds (N = 317)

| Threshold (kPa) | Overall agreement, % | $\kappa$ (95% CI) | VCTE+/2D-SWE-, n | VCTE-/2D-SWE+, n | Directional discordance proportion | McNemar *P* |
| --- | --- | --- | --- | --- | --- | --- |
| Lower (VCTE ≥ 8.0 / 2D-SWE ≥ 7.20) | 76.0 | .42 (.32 to .51) | 71 | 5 | .93 | < .001 |
| Upper (VCTE ≥ 10.0 / 2D-SWE ≥ 8.06) | 85.5 | .45 (.31 to .57) | 38 | 8 | .83 | < .001 |
| Sensitivity (VCTE ≥ 9.6 / 2D-SWE ≥ 8.06) | 84.5 | .44 (.30 to .56) | 42 | 7 | .86 | < .001 |

The symbols + and - indicate values at or above and below the corresponding method-specific threshold, respectively. Directional discordance proportion is the number of VCTE+/2D-SWE- pairs divided by all discordant pairs; 0.50 indicates symmetric discordance. Analyses used the unrounded published 2D-SWE thresholds of 7.204 and 8.060 kPa; threshold values are displayed to 2 decimal places. VCTE was used as an operational reference rather than a histologic gold standard.

### Prespecified Robustness and Exploratory Analyses

Excluding measurements within borderline zones modestly increased κ but did not remove discordance asymmetry (Supplementary Table S3). Across alternative published frameworks, agreement remained slight to moderate (κ, 0.22–0.51). Conventional or higher 2D-SWE thresholds predominantly produced VCTE-positive/2D-SWE-negative discordance, whereas very low 2D-SWE thresholds reversed its direction, indicating that disagreement was not attributable to a single cut-off (Supplementary Table S4).

Agreement worsened descriptively across increasing BMI strata: mean bias increased from 0.93 to 2.03 to 2.94 kPa, while the ICC decreased from 0.488 to 0.378 to 0.334. The BMI-stratified findings are shown in Supplementary Figure S5. XL-probe examinations had wider limits of agreement than M-probe examinations, and E9 examinations had a higher mean bias and geometric mean ratio than E10 examinations, partly reflecting the higher-stiffness distribution examined using E9 (Supplementary Tables S5–S6; Supplementary Figure S1).

Mean stiffness explained most of the variation in both absolute (R²=0.73) and signed discrepancy (R²=0.70). BMI, waist circumference, and controlled attenuation parameter showed weaker univariable associations, each explaining approximately 2%–5% of the variation; in multivariable models, waist circumference, selected to represent adiposity, and controlled attenuation parameter were not independently associated. Measurement-quality indices were not associated with either the magnitude or direction of disagreement, and stricter reliability restrictions did not eliminate the wide limits of agreement (Supplementary Tables S2 and S8–S9; Supplementary Figure S2). Signed-discrepancy results confirmed that increasing disagreement at higher stiffness occurred predominantly in the direction of higher VCTE values (Supplementary Table S15).

### Post-Hoc Analyses: Deming leverage sensitivity and operational threshold recalibration

The primary Deming slope was sensitive to high-stiffness, large-discrepancy observations, decreasing from 3.40 to 1.86 after excluding pairs with an absolute difference >5 kPa. Nevertheless, the slope remained above unity, with confidence intervals excluding identity, across all sensitivity scenarios (Supplementary Table S10; Supplementary Figure S3).

Continuous 2D-SWE discriminated the VCTE ≥8.0- and ≥10.0-kPa labels with areas under the curve of 0.810 (95% CI, 0.76–0.86) and 0.836 (95% CI, 0.77–0.90), respectively. Youden-derived 2D-SWE thresholds were 5.90 kPa (bootstrap 95% CI, 5.54–6.75) and 6.42 kPa (5.78– 7.15), substantially below the published comparators; internal validation showed limited optimism (Supplementary Table S11; Supplementary Figure S4). Alternative operating points and subgroup-specific candidates are reported in Supplementary Tables S12 and S14. Recalibration changed lower-threshold discordance from 71 versus 5 patients to 37 versus 39, leaving overall agreement unchanged at 76.0%. At the upper threshold, discordance changed from 38 versus 8 to 19 versus 32, while agreement decreased from 85.5% to 83.9% (Figure 3; Supplementary Table S13). Bootstrap analyses confirmed the reduction in directional imbalance but not an improvement in overall agreement (Supplementary Table S7). Thus, recalibration redistributed the direction of discordance without reducing total patient-level disagreement.

**Figure 3.**
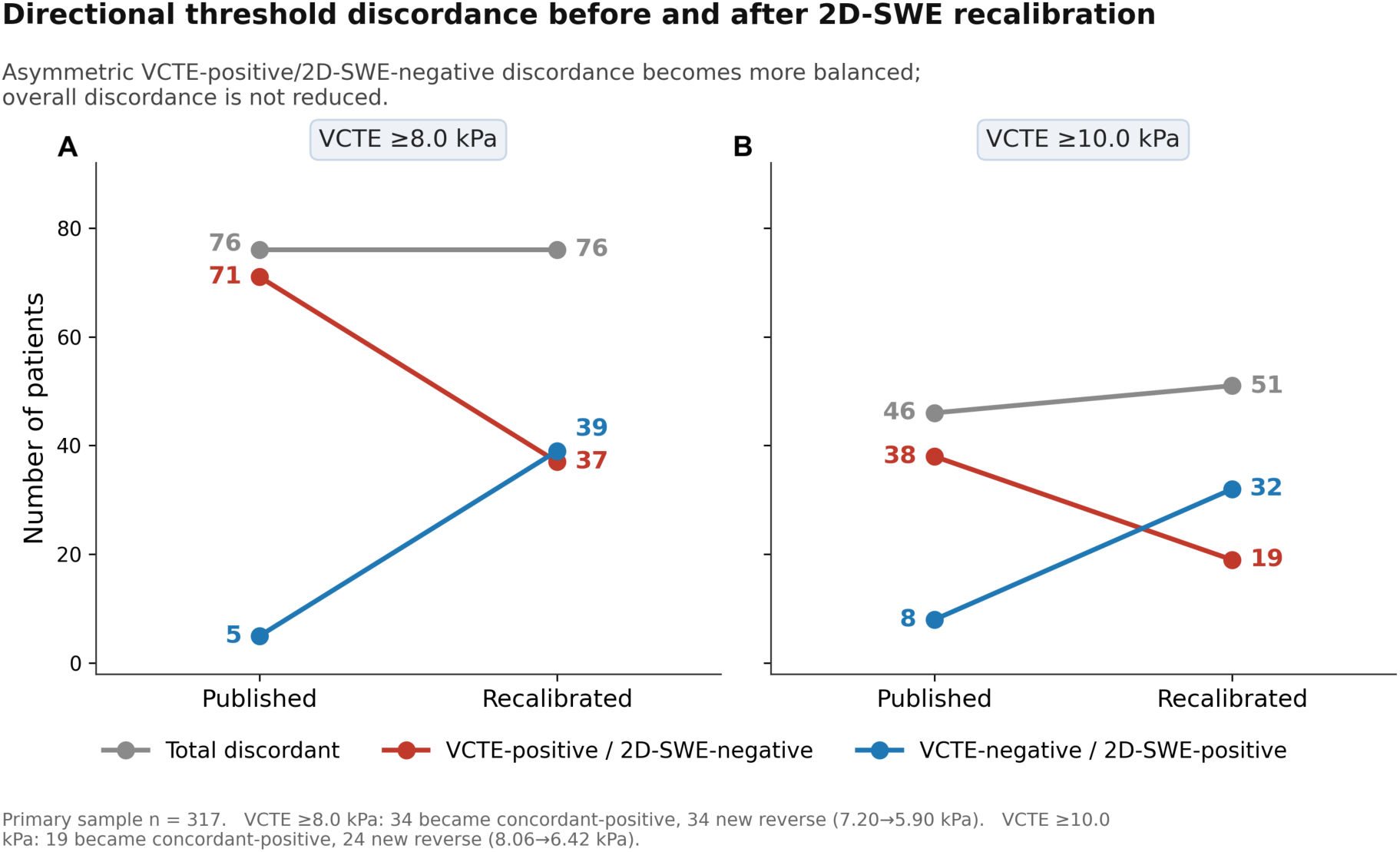
Directional threshold discordance before and after post hoc 2D-SWE recalibration. Published and recalibrated 2D-SWE thresholds are compared with the lower and upper VCTE decision thresholds. VCTE was used as an operational reference; recalibrated values are exploratory harmonization thresholds rather than diagnostic cut-offs.

## Discussion

In this same-day paired retrospective study of predominantly severely obese patients with MASLD, VCTE and GE 2D-SWE did not provide interchangeable liver stiffness measurements at the individual-patient level. VCTE values averaged 23% higher, but wide 95% ratio limits of agreement (0.64–2.40) showed substantial variation between individual patients. Threshold discordance was common and strongly directional: approximately three in five VCTE-positive patients were below the corresponding published 2D-SWE comparator at both thresholds, whereas reverse discordance was uncommon. Agreement also worsened descriptively across BMI strata. Lowering the 2D-SWE thresholds reduced the original directional imbalance but increased reverse discordance, leaving overall agreement unchanged or lower. Thus, neither an average numerical relationship nor adjustment of decision thresholds restores patient-level interchangeability between the two methods.

The central clinical question of the study was whether the two methods provide sufficiently similar numerical results in the same patient. They did not, as the wide limits of agreement on both the absolute and ratio scales preclude reliable patient-specific conversion of a 2D-SWE value into a corresponding VCTE value. The increasing difference at higher stiffness is particularly relevant because modality-related apparent changes were greatest in patients with higher LSM values. This distinction between association and agreement is also illustrated by previous comparisons using other 2D-SWE platforms. Studies using Supersonic Imagine and Canon systems reported strong correlations with VCTE (Pearson r = 0.882 and 0.932, respectively),^8, 9^ yet their Bland–Altman limits of agreement were wide, and both demonstrated increasing divergence or relatively lower 2D-SWE values in the higher stiffness range. Although the methods may therefore rank patients similarly, correlation does not establish that paired measurements are numerically equivalent. In our cohort, both Deming and Passing– Bablok regression were incompatible with the line of identity, further supporting a systematic difference in scale. The primary Deming slope was influenced by large-discrepancy observations, decreasing from 3.40 to 1.86 after exclusion of the 37 pairs differing by >5 kPa; however, it remained significantly above unity across sensitivity analyses. Likewise, the high AUROCs reported in previous studies demonstrate that 2D-SWE can discriminate between VCTE-defined categories,^8, 9^ but discrimination does not establish numerical agreement, calibration, or interchangeability for longitudinal measurements in an individual patient.

At the prespecified binary clinical decision thresholds, using a different modality frequently changed device-derived status. At both thresholds, approximately three in five VCTE-positive patients were classified below threshold by 2D-SWE, whereas reverse discordance was uncommon. Overall agreement and κ can obscure this directional imbalance because they combine discordant directions and are strongly influenced by concordant below-threshold pairs. Because VCTE was an operational reference rather than diagnostic truth, discordances cannot be interpreted as false-positive VCTE or false-negative 2D-SWE results.

The clinical implications extend beyond baseline risk classification. Longitudinal VCTE studies have shown that progression across 10 kPa and regression below it are associated with corresponding changes in liver-related-event risk, while percentage changes among patients starting at ≥8 or ≥10 kPa also carry prognostic information.^19, 20^ In our cohort, 37 patients (11.7%) had same-day intermethod differences >5 kPa, with VCTE higher in every case, and 25 crossed at least one prespecified decision threshold. A recent 16-center longitudinal study additionally found that a ≥5-kPa increase in serial VCTE was associated with increased liver-related-event risk in patients with MASLD and baseline LSM of 10–20 kPa.^23^ Although same-day intermethod differences cannot be equated with genuine longitudinal disease progression or regression, switching from 2D-SWE to VCTE could simulate a clinically consequential increase, whereas switching in the opposite direction could simulate improvement. Patients assessed initially with one modality should therefore preferably undergo serial measurements using the same modality and platform. If switching is unavoidable, the new measurement should not be interpreted as a direct continuation of the previous numerical series, and establishing a new method-specific baseline may be appropriate. These findings argue against assuming that VCTE and 2D-SWE are interchangeable in clinical practice or research.

Could the disagreement be resolved by applying different decision thresholds? Only partly. Across alternative published threshold frameworks, agreement remained limited (κ 0.22–0.51), and conventional or higher 2D-SWE cut-offs consistently produced predominantly VCTE-positive/2D-SWE-negative discordance. Excluding measurements close to either method-specific cut-off modestly improved κ but did not remove the directional asymmetry, indicating that disagreement was not confined to borderline observations. Conversely, applying very low 2D-SWE thresholds reversed the direction of discordance rather than eliminating it (Supplementary Tables S3–S4). Consistent with these findings, post hoc VCTE-referenced recalibration produced substantially lower 2D-SWE thresholds of 5.90 and 6.42 kPa, confirming that the published comparators were too high relative to the VCTE-referenced operating range in this cohort. Recalibration reduced VCTE-positive/2D-SWE-negative discordance from 71 to 37 patients at the lower threshold and from 38 to 19 at the upper threshold, but increased reverse discordance from 5 to 39 and from 8 to 32, respectively. Consequently, total disagreement was unchanged at the lower threshold and increased slightly at the upper threshold. Recalibration therefore rebalanced the direction of classification discordance without improving overall patient-level agreement. Adjusting a decision boundary cannot correct the underlying numerical non-equivalence or wide limits of agreement between the methods. Because the recalibrated values were derived and internally validated using VCTE labels as an operational reference, without histologic confirmation or external validation, they should be regarded as exploratory harmonization candidates rather than new diagnostic cut-offs for fibrosis or a clinical solution to intermethod disagreement.

The magnitude and direction of disagreement provide complementary clinical information. Absolute-discrepancy models address which patients are most likely to experience a large numerical change when the modality is switched, whereas signed-discrepancy models indicate the expected direction of that change. Higher mean stiffness was by far the dominant correlate of both outcomes, explaining 73% of the variation in absolute discrepancy and 70% of the variation in signed discrepancy. BMI, waist circumference, and CAP showed substantially weaker univariable associations, each explaining only approximately 2%–5% of the variation; in the adjusted models, waist circumference, used to represent adiposity, and CAP were not independently associated with discrepancy. Because mean stiffness was calculated from both modalities, these associations should be interpreted as describing where disagreement was greatest rather than as a prospective prediction rule available before switching methods. Nevertheless, agreement and threshold concordance worsened descriptively across BMI strata. Thus, patients in the higher stiffness range appear most vulnerable to clinically important modality-related changes, while severe obesity identifies a clinically relevant context in which agreement is poorer but should not be interpreted as an independently established cause.

Although VCTE and 2D-SWE both infer liver stiffness from shear-wave propagation, they differ in how the shear waves are generated and tracked. VCTE uses a controlled external mechanical vibration with fixed, non–image-guided sampling geometry, whereas 2D-SWE uses acoustic radiation force and permits B-mode-guided placement of a local measurement ROI.^24^ In this study, the reported 2D-SWE value was the median of 12 such acquisitions. Differences in shear-wave frequency and bandwidth, acquisition depth, sampling geometry and effective measurement volume, signal attenuation, quality filtering, and vendor-specific processing may therefore contribute to the observed difference in scale. Body habitus may further affect the acoustic window, signal transmission and attenuation, and measurement depth.^24^

A possible upper-range limitation was suggested by several highly discrepant observations in which VCTE ranged from 25.6 to 37.0 kPa while 2D-SWE remained between 11.3 and 13.6 kPa; no 2D-SWE value exceeded 13.62 kPa in the primary analysis population. Parallel interplatform evidence supports the plausibility of upper-range compression: in a same-day study using the LOGIQ E10, GE 2D-SWE values became substantially lower than Supersonic Imagine values above 13 kPa, with the greatest divergence above 17 kPa.^25^ This external evidence supports vendor-specific compression of higher values but does not establish a fixed technical ceiling. Accordingly, the pattern in our cohort is best described as possible high-range compression or a cohort-specific apparent ceiling among reliably obtained 2D-SWE measurements. The small number of highly discrepant observations and absence of an independent reference standard preclude distinguishing 2D-SWE underestimation from VCTE overestimation, tissue heterogeneity, or other acquisition-related effects. These explanations therefore remain hypotheses because the study was not designed to isolate any individual technical mechanism.

Previous comparisons in NAFLD or MASLD do not necessarily conflict with the present findings because most addressed diagnostic discrimination against histologic fibrosis rather than patient-level numerical agreement between modalities. In a biopsy-referenced cohort enriched for advanced fibrosis, Cassinotto et al. found similar performance of SSI 2D-SWE and VCTE within multistep diagnostic strategies and showed that a common 8–10-kPa gray zone could support clinically useful triage.^10^ That conclusion is appropriate for the evaluated SSI-based diagnostic algorithm but does not establish that paired measurements are numerically equivalent, particularly because agreement analyses and directional reclassification were not reported and the authors explicitly limited their findings to the SSI platform. Similarly, Sharpton et al. found comparable AUROCs for diagnosing significant fibrosis, advanced fibrosis, and cirrhosis with SSI 2D-SWE and VCTE.^11^ Their stage-specific measurements nevertheless showed increasing separation at higher fibrosis stages, with median VCTE and 2D-SWE values of 24.2 and 12.4 kPa, respectively, in cirrhosis. They also found poorer 2D-SWE performance in the highest BMI quartile. Thus, their findings support the diagnostic utility of both methods while also illustrating that comparable discrimination can coexist with substantial numerical divergence.

The most directly relevant biopsy-referenced comparison used GE LOGIQ Fortis 2D-SWE in patients with MASLD and found diagnostic performance comparable to VCTE for F≥1 and F≥2.^12^ However, intermethod correlation weakened at higher BMI, and mean VCTE values exceeded 2D-SWE values at F3 and F4. Only 11 patients had F3–F4 fibrosis, and correlation within this small subgroup did not establish agreement; consequently, the authors’ suggestion of potential interchangeability for monitoring advanced fibrosis requires confirmation by dedicated agreement and longitudinal analyses. In a larger non-biopsy study using SSI, Karagiannakis et al. reported strong correlation, good categorical agreement, and excellent reproduction of VCTE-defined advanced-fibrosis and cirrhosis classifications after optimization of 2D-SWE thresholds.^13^ These results demonstrate that 2D-SWE can reproduce selected VCTE-derived categories, but not that the numerical measurements can be continued interchangeably within an individual patient.

Two large method-comparison studies in mixed-etiology populations provide additional context. Cassinotto et al. reported strong correlation and favorable reproduction of VCTE-defined cACLD categories with SSI 2D-SWE, but only 28% of participants had NAFLD, median BMI was 27 kg/m², and VCTE was performed exclusively with the M probe.^8^ Despite the favorable categorical performance, their Bland–Altman analysis showed wide patient-level variation and relatively lower 2D-SWE values at higher stiffness. Ronot et al. similarly found that Canon 2D-SWE correlated strongly with VCTE and reproduced most VCTE-derived categories, while progressively underestimating higher VCTE measurements.^9^ That cohort was predominantly composed of patients with viral hepatitis, with only 3.4% having NAFLD and fewer than 10% having obesity. Importantly, the authors recommended that longitudinal examinations should use the same system to reduce misclassification. These studies therefore support rather than contradict the present findings: categorical risk stratification may remain clinically useful after platform-specific threshold adjustment, while wide individual variation and proportional bias preclude assuming numerical interchangeability.

Collectively, the literature supports the clinical validity of both modalities but also demonstrates that 2D-SWE is not a uniform technology and that findings obtained with one manufacturer cannot automatically be generalized to another. The present study extends this literature by evaluating same-day, patient-level numerical agreement and directional threshold reclassification with GE LOGIQ E9/E10 2D-SWE in a predominantly severely obese MASLD population. The predominance of severe obesity in our cohort should not be viewed solely as an atypical sampling feature. In a real-world US health-system cohort of 32,900 patients with MASLD, 92.6% were overweight or obese and 68.2% had BMI ≥30 kg/m²; the relationship between BMI and outcomes was J-shaped, with very severe obesity associated with the greatest risks of hepatic decompensation and all-cause mortality during a mean follow-up of 5.5 years.^26^ Recent epidemiologic evidence similarly shows that MASLD prevalence rises markedly with increasing obesity class and emphasizes continued assessment of liver status during obesity-directed or liver-targeted therapy.^27^ Our cohort therefore represents an enriched tertiary-care phenotype rather than the complete MASLD spectrum, but one that is increasingly common, clinically consequential, and particularly likely to undergo repeated LSM during treatment and follow-up. The findings are most directly generalizable to severely obese patients managed in tertiary, metabolic, or bariatric pathways, in whom maintaining consistency of elastography modality may be especially important. Accordingly, published platform-specific thresholds may remain diagnostically appropriate against their original reference standard while still being too high relative to the VCTE-referenced operating range in this cohort.

Several features strengthen this study. VCTE and 2D-SWE were performed on the same day after standardized prolonged fasting, minimizing biological and temporal variation between paired measurements. Examinations were undertaken by separate experienced operators who were not informed of the other modality’s result, using standardized acquisition procedures and rigorous reliability criteria. The study evaluated a clinically important population of patients with MASLD and severe obesity who are frequently assessed in tertiary, metabolic, and bariatric pathways. Continuous and categorical agreement analyses were prespecified, both directions of threshold discordance were reported, and the main findings were examined across extensive subgroup, device, and leverage-sensitivity analyses. Prespecified and post hoc analyses were also clearly distinguished, including transparent identification of the recalibration analyses as exploratory.

These strengths should be considered alongside several limitations. The retrospective, single-center design limits generalizability, although it allowed paired examinations to be acquired under a consistent clinical protocol. The predominantly female and severely obese cohort does not represent the complete MASLD spectrum; the findings are most directly applicable to similar tertiary-care populations and should not be extrapolated automatically to lean MASLD or other disease etiologies. Because inclusion required valid paired examinations, the study estimates agreement conditional on successful measurement and cannot compare the feasibility or failure rates of the two modalities. No biopsy or magnetic resonance elastography reference was available; consequently, VCTE served only as an operational reference for categorical analyses, and discordant results cannot be assigned as errors of either modality. An independent fibrosis reference was not required to evaluate whether paired measurements agreed, but its absence prevents conclusions about which method more accurately reflects fibrosis. The 2D-SWE findings are specific to the GE LOGIQ E9/E10 implementation and should not be generalized to other manufacturers. Relatively few patients had very high stiffness, and observations with large intermethod differences influenced the regression slopes; these slopes should therefore not be interpreted as conversion factors, although proportional bias persisted in sensitivity analyses. Finally, the cross-sectional design did not directly evaluate the longitudinal consequences of switching modalities, and the post hoc recalibrated thresholds, despite internal validation, require external validation and should not be considered diagnostic cut-offs.

## Conclusion

In this real-world cohort of predominantly severely obese patients with MASLD, VCTE and GE LOGIQ 2D-SWE did not provide interchangeable patient-level liver stiffness measurements. VCTE values were generally higher, divergence increased at higher stiffness, and approximately three in five VCTE-positive patients fell below the corresponding 2D-SWE decision threshold. Lower recalibrated cut-offs redistributed the direction of discordance without reducing its overall extent, demonstrating that threshold adjustment cannot resolve the underlying numerical non-equivalence. Serial LSM should therefore be performed using the same elastography modality and, when possible, the same platform. If switching is unavoidable, establishing a new method-specific baseline is preferable to interpreting the new measurement as a direct continuation of the previous series. These findings do not establish which modality more accurately reflects fibrosis, but demonstrate that changing modalities may create an apparent change that does not represent true disease progression or regression.

## Data Availability

All data produced in the present study are available upon reasonable request to the authors

## Author contributions

V.I.: Conceptualization, Supervision, Resources, Methodology, Writing—Original Draft, Writing—Review and Editing; A.G.: Investigation, Data Curation, Software, Formal Analysis, Writing—Review and Editing; M.I.: Visualization, Investigation, Writing—Review and Editing.

## Data Transparency Statement

The statistical analysis plan is publicly archived on the Open Science Framework (https://doi.org/10.17605/OSF.IO/E95D8), and its post-hoc amendment is archived separately (https://doi.org/10.17605/OSF.IO/E84R7); both documents are also provided as Supplementary Material. R scripts and de-identified patient-level data supporting the findings are available from the corresponding author on reasonable request, subject to institutional approval and a data-sharing agreement, and in accordance with applicable data-protection regulations.

## References

1. Feng G, Targher G, Byrne CD, et al. Global burden of metabolic dysfunction-associated steatotic liver disease, 2010 to 2021. JHEP Rep. 2025;7(3):101271.

2. Golabi P, Isakov V, Younossi ZM. Nonalcoholic Fatty Liver Disease: Disease Burden and Disease Awareness. Clin Liver Dis. 2023;27(2):173–86.

3. Dulai PS, Singh S, Patel J, et al. Increased risk of mortality by fibrosis stage in nonalcoholic fatty liver disease: Systematic review and meta-analysis. Hepatology. 2017;65(5):1557–65.

4. Taylor RS, Taylor RJ, Bayliss S, et al. Association Between Fibrosis Stage and Outcomes of Patients With Nonalcoholic Fatty Liver Disease: A Systematic Review and Meta-Analysis. Gastroenterology. 2020;158(6):1611–25 e12.

5. Rinella ME, Neuschwander-Tetri BA, Siddiqui MS, et al. AASLD Practice Guidance on the clinical assessment and management of nonalcoholic fatty liver disease. Hepatology. 2023;77(5):1797–835.

6. Tacke F, Horn P, Wai-Sun Wong V, et al. EASL-EASD-EASO Clinical Practice Guidelines on the management of metabolic dysfunction-associated steatotic liver disease (MASLD). J Hepatol. 2024;81(3):492–542.

7. Castera L, Rinella ME, Tsochatzis EA. Noninvasive Assessment of Liver Fibrosis. N Engl J Med. 2025;393(17):1715–29.

8. Cassinotto C, Lapuyade B, Guiu B, et al. Agreement Between 2-Dimensional Shear Wave and Transient Elastography Values for Diagnosis of Advanced Chronic Liver Disease. Clin Gastroenterol Hepatol. 2020;18(13):2971–9 e3.

9. Ronot M, Ferraioli G, Muller HP, et al. Comparison of liver stiffness measurements by a 2D-shear wave technique and transient elastography: results from a European prospective multi-centre study. Eur Radiol. 2021;31(3):1578–87.

10. Cassinotto C, Boursier J, Paisant A, et al. Transient Versus Two-Dimensional Shear-Wave Elastography in a Multistep Strategy to Detect Advanced Fibrosis in NAFLD. Hepatology. 2021;73(6):2196–205.

11. Sharpton SR, Tamaki N, Bettencourt R, et al. Diagnostic accuracy of two-dimensional shear wave elastography and transient elastography in nonalcoholic fatty liver disease. Ther Adv Gastroenterol. 2021;14:17562848211050436.

12. Li Q, Gao Y, Wang D, et al. Prospective Comparison of 2D-Shear Wave Elastography and Vibration-Controlled Transient Elastography in Assessing Liver Fibrosis in Metabolic Dysfunction-Associated Steatotic Liver Disease. Acad Radiol. 2025;32(9):5100–11.

13. Karagiannakis DS, Markakis G, Lakiotaki D, et al. Comparing 2D-shear wave to transient elastography for the evaluation of liver fibrosis in nonalcoholic fatty liver disease. Eur J Gastroenterol Hepatol. 2022;34(9):961–6.

14. Kanwal F, Neuschwander-Tetri BA, Loomba R, et al. Metabolic dysfunction–associated steatotic liver disease: Update and impact of new nomenclature on the American Association for the Study of Liver Diseases practice guidance on nonalcoholic fatty liver disease. Hepatology. 2024;79(5):1212–9.

15. Boursier J, Zarski JP, de Ledinghen V, et al. Determination of reliability criteria for liver stiffness evaluation by transient elastography. Hepatology. 2013;57(3):1182–91.

16. General Electric Company. LOGIQ E10 Shear Wave Elastography: Abdominal Imaging. 2018 JB57257XX.

17. Furlan A, Tublin ME, Yu L, et al. Comparison of 2D Shear Wave Elastography, Transient Elastography, and MR Elastography for the Diagnosis of Fibrosis in Patients With Nonalcoholic Fatty Liver Disease. AJR Am J Roentgenol. 2020;214(1):W20–W6.

18. Mozes FE, Lee JA, Selvaraj EA, et al. Diagnostic accuracy of non-invasive tests for advanced fibrosis in patients with NAFLD: an individual patient data meta-analysis. Gut. 2022;71(5):1006–19.

19. Gawrieh S, Vilar-Gomez E, Wilson LA, et al. Increases and decreases in liver stiffness measurement are independently associated with the risk of liver-related events in NAFLD. J Hepatol. 2024;81(4):600–8.

20. Pennisi G, Infantino G, Di Maria G, et al. One-Year Changes in Liver Stiffness Measurement, but not in Alanine Aminotransferase and Controlled Attenuation Parameter, Predict Long-Term Liver Outcomes in Patients With Metabolic Dysfunction-Associated Steatotic Liver Disease. Clin Gastroenterol Hepatol. 2026:In Press.

21. Indre MG, Leucuta DC, Lupsor-Platon M, et al. Diagnostic accuracy of 2D-SWE ultrasound for liver fibrosis assessment in MASLD: A multilevel random effects model meta-analysis. Hepatology. 2025;82(2):454–69.

22. General Electric Company. 2D Shear Wave Elastography LOGIQ E9/E10/E10s. General Electric Company; 2020; Available from: https://gehealthcare-ultrasound.com/en/logiq-family/logiq-e10-series/download-liver-fibrosis/.

23. Jin X, Peng N, Lin H, et al. Prognostic significance of a 5 kPa absolute change in liver stiffness measurements (LSM) to predict liver-related events in patients with compensated advanced metabolic dysfunction-associated steatotic liver disease (MASLD). J Hepatol. 2026;84(Suppl 1):S589.

24. Kennedy P, Wagner M, Castera L, et al. Quantitative Elastography Methods in Liver Disease: Current Evidence and Future Directions. Radiology. 2018;286(3):738–63.

25. Cassinotto C, Anselme S, Jacq T, et al. Inter-platform Variability of Liver Elastography: Pairwise Comparisons of Four Devices. Ultrasound Med Biol. 2022;48(11):2258–66.

26. Behari J, Wang R, Luu HN, et al. Severe obesity is associated with worse outcomes than lean metabolic dysfunction-associated steatotic liver disease. Hepatol Commun. 2024;8(7):e0471.

27. Grocic M, Nguyen M, Sanyal AJ. Current Trends in the Epidemiology of Obesity and the Association Between Obesity and Metabolic Liver Disease (MASLD/MASH). Diabetes Obes Metab. 2026;28 Suppl 2(Suppl 2):3–18.

