## Supplementary Tables S1-S15 for "Switching Between VCTE and 2D-SWE Reclassifies Fibrosis Risk in Metabolic Dysfunction-Associated Steatotic Liver Disease"

#### Supplementary Table S1. Analysis Population and Data Completeness (N = 317)

| Variable | n available | n missing | Missing, % |
|---|---|---|---|
| Age | 317 | 0 | 0.0 |
| Sex | 317 | 0 | 0.0 |
| Body mass index | 317 | 0 | 0.0 |
| Waist circumference | 317 | 0 | 0.0 |
| Hip circumference | 317 | 0 | 0.0 |
| Waist-to-hip ratio | 317 | 0 | 0.0 |
| Alanine aminotransferase | 317 | 0 | 0.0 |
| VCTE probe (M/XL) | 317 | 0 | 0.0 |
| VCTE liver stiffness | 317 | 0 | 0.0 |
| VCTE IQR | 317 | 0 | 0.0 |
| VCTE IQR/median | 317 | 0 | 0.0 |
| Controlled attenuation parameter | 317 | 0 | 0.0 |
| CAP IQR | 317 | 0 | 0.0 |
| 2D-SWE liver stiffness | 317 | 0 | 0.0 |
| 2D-SWE IQR/median | 317 | 0 | 0.0 |

NOTE. All variables used in the analyses were complete; no imputation was performed and all analyses were complete-case (N = 317).

**Supplementary Table S2. Agreement Under Reliability (Measurement-Quality) Criteria**

| Quality criterion | n | Bias, kPa | Lower LoA, kPa | Upper LoA, kPa | ICC |
|---|---|---|---|---|---|
| Strict conventional ≤ 30%* (overrides Boursier "Also reliable") | 278 | 2.20 | −4.88 | 9.29 | .40 |
| Both IQR/median ≤ 25% | 193 | 1.97 | −5.28 | 9.22 | .44 |
| Both IQR/median ≤ 20% | 113 | 1.28 | −3.64 | 6.20 | .48 |
| VCTE ≤ 30% only | 278 | 2.20 | −4.88 | 9.29 | .40 |

Analyses restricted to measurements meeting each reliability criterion. Bias is the mean of (VCTE − 2D-SWE); IQR/median is the interquartile range divided by the median stiffness. * excludes Boursier-reliable examinations with IQR/median >30% and median LSM <7.1 kPa.

**Supplementary Table S3. Borderline-Zone Sensitivity Analysis at the Decision Thresholds**

| Threshold | Excluded zone | n excluded | n remaining | κ | McNemar *P* |
|---|---|---|---|---|---|
| Lower | ±10% | 66 | 251 | .57 | < .001 |
| Upper | ±10% | 55 | 262 | .52 | < .001 |
| Lower | ±15% | 117 | 200 | .63 | < .001 |
| Upper | ±15% | 84 | 233 | .58 | < .001 |
| Lower | ±0.5 kPa | 45 | 272 | .54 | < .001 |
| Upper | ±0.5 kPa | 30 | 287 | .49 | < .001 |
| Lower | ±1.0 kPa | 104 | 213 | .59 | < .001 |
| Upper | ±1.0 kPa | 63 | 254 | .53 | .002 |

Patients whose liver stiffness fell within the specified zone around each threshold were excluded and agreement re-estimated.

κ, Cohen kappa.

**Supplementary Table S4. Agreement Across Alternative Threshold Frameworks**

| Framework | n | Agreement, % | κ (95% CI) | VCTE+/2D-SWE−, n | VCTE−/2D-SWE+, n | McNemar *P* |
|---|---|---|---|---|---|---|
| Primary GE lower | 317 | 76.0 | .42 (.32 to .51) | 71 | 5 | < .001 |
| Primary GE upper | 317 | 85.5 | .45 (.31 to .57) | 38 | 8 | < .001 |
| All-vendor MASLD (2D-SWE F2) | 317 | 71.9 | .29 (.21 to .39) | 88 | 1 | < .001 |
| All-vendor MASLD (2D-SWE F3) | 317 | 82.3 | .22 (.11 to .36) | 52 | 4 | < .001 |
| GE white-paper (2D-SWE F2) | 317 | 71.0 | .26 (.18 to .36) | 91 | 1 | < .001 |
| GE white-paper (2D-SWE F3) | 317 | 82.3 | .22 (.11 to .36) | 52 | 4 | < .001 |
| Furlan significant (Youden 5.7) | 317 | 72.2 | .43 (.33 to .53) | 33 | 55 | .02 |
| Furlan significant (high-Sp 7.2) | 317 | 77.0 | .44 (.35 to .54) | 68 | 5 | < .001 |
| Furlan advanced (Youden 8.1) | 317 | 85.5 | .45 (.31 to .57) | 38 | 8 | < .001 |
| Furlan advanced (high-Sp 8.0) | 317 | 85.2 | .44 (.30 to .56) | 38 | 9 | < .001 |
| Furlan advanced (rule-out 6.4) | 317 | 83.0 | .51 (.40 to .63) | 19 | 35 | .03 |
| VCTE exact 9.6 vs GE upper | 317 | 84.5 | .44 (.30 to .56) | 42 | 7 | < .001 |
| VCTE cACLD 10 vs GE upper | 317 | 85.5 | .45 (.31 to .57) | 38 | 8 | < .001 |
| VCTE 15 vs GE upper | 317 | 92.1 | .43 (.24 to .62) | 3 | 22 | < .001 |
| VCTE 20 vs all-vendor F4 | 317 | 98.4 | .54 (.00 to .87) | 4 | 1 | .18 |
| VCTE 20 vs GE white-paper F4 | 317 | 98.4 | .54 (.00 to .87) | 4 | 1 | .18 |

Cut-off values (kPa) for each framework are given in the Methods and Supplementary Statistical Report. VCTE+ denotes at or above the VCTE threshold and 2D-SWE− below the paired 2D-SWE threshold. cACLD, compensated advanced chronic liver disease; CI, confidence interval; VCTE, vibration-controlled transient elastography; κ, Cohen kappa.

**Supplementary Table S5. Agreement Stratified by VCTE Probe**

| Probe | n | Bias, kPa | Lower LoA, kPa | Upper LoA, kPa | ICC | κ (lower) | κ (upper) |
|---|---|---|---|---|---|---|---|
| M | 80 | 1.31 | −3.04 | 5.66 | .39 | .40 | .33 |
| XL | 237 | 2.11 | −5.40 | 9.62 | .40 | .41 | .47 |

Bias is the mean of (VCTE − 2D-SWE). κ is reported at the lower and upper decision thresholds.

ICC, intraclass correlation coefficient; LoA, 95% limits of agreement; VCTE, vibration-controlled transient elastography; κ, Cohen kappa.

**Supplementary Table S6. Agreement Stratified by 2D-SWE Device (GE LOGIQ E9 vs E10)**

| Device | n | Bias, kPa | Lower LoA, kPa | Upper LoA, kPa | Geometric mean ratio | κ (lower) | κ (upper) |
|---|---|---|---|---|---|---|---|
| E9 | 216 | 2.26 | −4.87 | 9.38 | 1.28 | .38 | .45 |
| E10 | 102 | 1.20 | −4.90 | 7.30 | 1.14 | .46 | .33 |

Bias is the mean of (VCTE − 2D-SWE); the geometric mean ratio is VCTE/2D-SWE. κ is reported at the lower and upper decision thresholds. One patient was examined on both devices and contributes one measurement to each device stratum; the stratified totals therefore sum to 318, whereas the primary analysis used one measurement per patient (N = 317). LoA, 95% limits of agreement; κ, Cohen kappa.

**Supplementary Table S7. Change in Agreement After Recalibration (Bias-Corrected Accelerated Bootstrap, 2000 Resamples)**

| Metric | Estimate | 95% CI |
|---|---|---|
| VCTE ≥ 8.0 kPa | | |
| Δ Agreement (proportion) | .00 | −.04 to .06 |
| Δ κ | .07 | .01 to .18 |
| Δ VCTE+/2D-SWE− pairs (n) | −34 | −52 to −10 |
| Δ Directional discordance proportion | −.45 | −.64 to −.04 |
| VCTE ≥ 10.0 kPa | | |
| Δ Agreement (proportion) | −.02 | −.14 to .04 |
| Δ κ | .09 | −.07 to .22 |
| Δ VCTE+/2D-SWE− pairs (n) | −19 | −36 to −11 |
| Δ Directional discordance proportion | −.45 | −.77 to −.21 |
| VCTE ≥ 9.6 kPa (sensitivity) | | |
| Δ Agreement (proportion) | −.00 | −.12 to .04 |
| Δ κ | .12 | −.05 to .24 |
| Δ VCTE+/2D-SWE− pairs (n) | −21 | −39 to −13 |
| Δ Directional discordance proportion | −.44 | −.78 to −.26 |

Positive Δκ indicates improved chance-corrected agreement; Δ agreement near 0 indicates unchanged total concordance; negative Δ in VCTE+/2D-SWE− pairs and in the directional discordance proportion indicates removal of directional bias. 2D-SWE, two-dimensional shear-wave elastography; CI, confidence interval; VCTE, vibration-controlled transient elastography; κ, Cohen kappa.

**Supplementary Table S8. Univariable Predictors of the Absolute VCTE − 2D-SWE Difference**

| Predictor | β, kPa | 95% CI | *P* value | BH q value | $R^2$ |
|---|---|---|---|---|---|
| Age, per year | 0.04 | 0.00 to 0.07 | .03 | .067 | .02 |
| Female sex | 0.74 | −0.03 to 1.52 | .06 | .103 | .01 |
| BMI, per kg/m$^2$ | 0.07 | 0.03 to 0.12 | .002 | .009 | .03 |
| Waist circumference, per cm | 0.05 | 0.02 to 0.07 | < .001 | .001 | .04 |
| CAP, per dB/m | 0.01 | 0.00 to 0.02 | .005 | .017 | .02 |
| log(ALT) | 0.57 | −0.03 to 1.16 | .06 | .103 | .01 |
| XL probe | 0.77 | −0.03 to 1.57 | .06 | .103 | .01 |
| VCTE IQR/median, per percentage point | 0.01 | −0.03 to 0.04 | .76 | .803 | .00 |
| 2D-SWE IQR/median, per percentage point | 0.01 | −0.05 to 0.06 | .80 | .803 | .00 |
| LOGIQ E10 device | −0.86 | −1.61 to −0.12 | .02 | .065 | .02 |
| Mean LSM, per kPa | 0.92 | 0.86 to 0.99 | < .001 | < .001 | .73 |

Linear regression of the absolute stiffness difference on each predictor separately. BH q values are Benjamini–Hochberg false-discovery-rate-adjusted values from the prespecified pool of 41 exploratory tests. Reference categories were male sex, M probe, and LOGIQ E9. Positive coefficients indicate a larger absolute VCTE–2D-SWE difference. 2D-SWE, two-dimensional shear-wave elastography; ALT, alanine aminotransferase; BH, Benjamini–Hochberg; BMI, body mass index; CAP, controlled attenuation parameter; CI, confidence interval; IQR, interquartile range; LSM, liver stiffness measurement; $R^2$, coefficient of determination; VCTE, vibration-controlled transient elastography.

**Supplementary Table S9. Multivariable Predictors of the Absolute VCTE − 2D-SWE Difference**

| Predictor | Adjusted β, kPa | 95% CI | *P* value | BH q value |
|---|---|---|---|---|
| Intercept | −1.02 | −3.34 to 1.30 | .39 | — |
| Age, per year | −0.02 | −0.04 to −0.00 | .03 | .067 |
| Female sex | 0.40 | 0.00 to 0.79 | .05 | .102 |
| Waist circumference, per cm | −0.01 | −0.03 to 0.00 | .16 | .224 |
| CAP, per dB/m | 0.00 | −0.01 to 0.01 | .79 | .803 |
| log(ALT) | −0.48 | −1.00 to 0.03 | .06 | .103 |
| XL probe | −0.09 | −0.55 to 0.37 | .69 | .765 |
| LOGIQ E10 device | 0.44 | 0.06 to 0.81 | .02 | .065 |
| Mean LSM, per kPa | 0.99 | 0.85 to 1.12 | < .001 | < .001 |

Multivariable linear regression with all listed covariates entered simultaneously. BH q values are Benjamini–Hochberg false-discovery-rate-adjusted values from the prespecified pool of 41 exploratory tests; the intercept was not part of the adjusted pool. Reference categories were male sex, M probe, and LOGIQ E9. ALT, alanine aminotransferase; BH, Benjamini–Hochberg; CAP, controlled attenuation parameter; CI, confidence interval; LSM, liver stiffness measurement; NA, not applicable; VCTE, vibration-controlled transient elastography.

### Supplementary Table S10. Deming Regression Leverage Sensitivity Analyses

| Analysis | n | λ | Deming slope (95% CI) | Deming intercept (95% CI) |
|---|---|---|---|---|
| Primary (full sample, λ = 1) | 317 | 1.00 | 3.40 (2.94 to 3.86) | −12.05 (−14.56 to −9.55) |
| S1: exclude \|VCTE − 2D-SWE\| > 5 kPa (n = 37 excluded) | 280 | 1.00 | 1.86 (1.57 to 2.16) | −3.91 (−5.50 to −2.31) |
| S2: exclude Cook's D > 4/n (n = 17 excluded) | 300 | 1.00 | 2.80 (2.39 to 3.22) | −8.62 (−10.87 to −6.38) |
| S3: exclude VCTE or 2D-SWE LSM above its 99th percentile (n = 5 excluded) | 312 | 1.00 | 3.29 (2.57 to 4.01) | −11.43 (−15.35 to −7.51) |
| S4: winsorized at the 2.5th and 97.5th percentiles | 317 | 1.00 | 2.98 (2.53 to 3.43) | −9.71 (−12.15 to −7.27) |
| S5: mean LSM ≤ 12 kPa restriction (n = 14 excluded) | 303 | 1.00 | 2.70 (2.21 to 3.20) | −8.20 (−10.85 to −5.54) |
| S7a: full sample, λ = 0.5 | 317 | 0.50 | 3.18 (2.68 to 3.67) | −10.76 (−13.44 to −8.08) |
| S7b: full sample, λ = 2 | 317 | 2.00 | 3.52 (3.08 to 3.97) | −12.79 (−15.24 to −10.34) |

λ is the assumed ratio of squared measurement errors of the reference method to the test method. With 2D-SWE specified as the reference method and VCTE as the test method, $\lambda = \sigma^2_{error,2D\text{-}SWE}/\sigma^2_{error,VCTE}$. The primary analysis assumed equal error variances (λ = 1.0); λ = 0.5 and λ = 2.0 were examined as sensitivity analyses. In a separate leave-one-out analysis (S6), the Deming slope ranged from 3.27 to 3.46 across 317 refits, indicating that no single observation accounted for the primary slope. The prespecified full-sample Passing–Bablok result is reported in Table 2. 2D-SWE, two-dimensional shear-wave elastography; CI, confidence interval; LSM, liver stiffness measurement; VCTE, vibration-controlled transient elastography.

### Supplementary Table S11. Internal Validation of Recalibrated 2D-SWE Thresholds (Recalibration Analysis R1)

| VCTE target | n | Events | AUROC (95% CI) | Youden J (apparent) | Youden J (optimism-corrected) | Recalibrated cut, kPa (95% CI) |
|---|---|---|---|---|---|---|
| VCTE ≥ 8.0 kPa | 317 | 117 | .81 (.76 to .86) | .49 | .46 | 5.89 (5.54 to 6.75) |
| VCTE ≥ 10.0 kPa | 317 | 63 | .84 (.77 to .90) | .57 | .54 | 6.42 (5.78 to 7.15) |
| VCTE ≥ 9.6 kPa (sensitivity) | 317 | 68 | .84 (.78 to .90) | .57 | .54 | 6.42 (5.78 to 6.88) |
| VCTE ≥ 15 kPa (descriptive) | 317 | 14 | .94 (.88 to .99) | NA | NA | NA |
| VCTE ≥ 20 kPa (descriptive) | 317 | 7 | .92 (.81 to 1.00) | NA | NA | NA |

AUROC and Youden J were internally validated by optimism-corrected bootstrapping (500 resamples). The optimism-corrected Youden J is the apparent value minus the mean bootstrap optimism. Recalibrated cut-offs (Youden index) are given with paired bootstrap 95% CIs; descriptive targets below the events-per-variable floor are shown without recalibrated cut-offs. 2D-SWE, two-dimensional shear-wave elastography; AUROC, area under the receiver-operating-characteristic curve; CI, confidence interval; NA, not applicable; VCTE, vibration-controlled transient elastography.

**Supplementary Table S12. Candidate 2D-SWE Cut-offs by Operating-Point Criterion (Recalibration Analysis R2)**

| Criterion | 2D-SWE cut, kPa | Sensitivity | Specificity |
|---|---|---|---|
| VCTE ≥ 8.0 kPa | | | |
| Youden | 5.89 | .68 | .81 |
| Equal Se/Sp | 5.64 | .73 | .72 |
| High-Se (≥ .90) | 4.79 | .91 | .42 |
| High-Sp (≥ .90) | 6.30 | .56 | .91 |
| VCTE ≥ 10.0 kPa | | | |
| Youden | 6.42 | .70 | .87 |
| Equal Se/Sp | 6.04 | .75 | .75 |
| High-Se (≥ .90) | 4.91 | .90 | .41 |
| High-Sp (≥ .90) | 6.71 | .65 | .90 |
| VCTE ≥ 9.6 kPa (sensitivity) | | | |
| Youden | 6.42 | .69 | .88 |
| Equal Se/Sp | 6.02 | .76 | .76 |
| High-Se (≥ .90) | 4.91 | .91 | .41 |
| High-Sp (≥ .90) | 6.66 | .63 | .90 |

Candidate 2D-SWE thresholds re-derived against the VCTE decision-threshold label under alternative operating-point criteria. Sensitivity and specificity are relative to the VCTE operational reference. 2D-SWE, two-dimensional shear-wave elastography; Se, sensitivity; Sp, specificity; VCTE, vibration-controlled transient elastography.

**Supplementary Table S13. Performance of Published Versus Recalibrated 2D-SWE Thresholds (Recalibration Analysis R3)**

| Cut-off | 2D-SWE cut, kPa | Agreement, % | κ | Sensitivity | Specificity | VCTE+/2D-SWE−, n | VCTE−/2D-SWE+, n | McNemar *P* |
|---|---|---|---|---|---|---|---|---|
| VCTE ≥ 8.0 kPa | | | | | | | | |
| Published | 7.20 | 76.0 | .42 | .39 | .97 | 71 | 5 | < .001 |
| Recalibrated (Youden) | 5.89 | 76.0 | .49 | .68 | .81 | 37 | 39 | .82 |
| VCTE ≥ 10.0 kPa | | | | | | | | |
| Published | 8.06 | 85.5 | .45 | .40 | .97 | 38 | 8 | < .001 |
| Recalibrated (Youden) | 6.42 | 83.9 | .53 | .70 | .87 | 19 | 32 | .07 |
| VCTE ≥ 9.6 kPa (sensitivity) | | | | | | | | |
| Published | 8.06 | 84.5 | .44 | .38 | .97 | 42 | 7 | < .001 |
| Recalibrated (Youden) | 6.42 | 84.2 | .55 | .69 | .88 | 21 | 29 | .26 |

Sensitivity and specificity are relative to the VCTE operational reference. Recalibrated thresholds are candidate harmonisation values requiring external validation. VCTE+ denotes at or above the VCTE threshold and 2D-SWE− below the paired 2D-SWE threshold. 2D-SWE, two-dimensional shear-wave elastography; VCTE, vibration-controlled transient elastography; κ, Cohen kappa.

**Supplementary Table S14. Subgroup-Specific Recalibrated 2D-SWE Thresholds (Recalibration Analysis R4)**

| Subgroup | n | Events | Recalibrated cut, kPa | AUROC | Comment |
|---|---|---|---|---|---|
| VCTE ≥ 8.0 kPa | | | | | |
| Body mass index < 35 $kg/m^2$ | 122 | 20 | 5.61 | .84 | |
| Body mass index 35–39.9 $kg/m^2$ | 89 | 36 | 5.87 | .78 | |
| Body mass index ≥ 40 $kg/m^2$ | 106 | 61 | 5.92 | .75 | |
| Probe M | 80 | 22 | 5.88 | .79 | |
| Probe XL | 237 | 95 | 6.32 | .81 | |
| Device E9 | 216 | 98 | 6.51 | .79 | |
| Device E10 | 101 | 19 | NA | NA | Below event floor; descriptive only |
| VCTE ≥ 10.0 kPa | | | | | |
| Body mass index < 35 $kg/m^2$ | 122 | 12 | NA | NA | Below event floor; descriptive only |
| Body mass index 35–39.9 $kg/m^2$ | 89 | 17 | NA | NA | Below event floor; descriptive only |
| Body mass index ≥ 40 $kg/m^2$ | 106 | 34 | 6.41 | .83 | |
| Probe M | 80 | 13 | NA | NA | Below event floor; descriptive only |
| Probe XL | 237 | 50 | 6.58 | .85 | |
| Device E9 | 216 | 53 | 6.58 | .82 | |
| Device E10 | 101 | 10 | NA | NA | Below event floor; descriptive only |

Subgroup recalibration is exploratory; subgroups below the minimum events-per-variable floor are marked descriptive only and shown without a recalibrated cut-off.

2D-SWE, two-dimensional shear-wave elastography; AUROC, area under the receiver-operating-characteristic curve; NA, not applicable; VCTE, vibration-controlled transient elastography.

**Supplementary Table S15. Predictors of the Signed VCTE − 2D-SWE Difference (n = 317)**

**Panel A. Univariable models**

| Predictor | β, kPa | 95% CI | *P* value | BH q value | $R^2$ |
|---|---|---|---|---|---|
| Age, per year | 0.025 | −0.010 to 0.059 | .158 | .224 | .006 |
| Female sex | 0.613 | −0.252 to 1.477 | .164 | .224 | .006 |
| BMI, per kg/m$^2$ | 0.097 | 0.045 to 0.149 | < .001 | .002 | .041 |
| Waist circumference, per cm | 0.054 | 0.027 to 0.080 | < .001 | < .001 | .049 |
| CAP, per dB/m | 0.018 | 0.007 to 0.028 | .001 | .008 | .032 |
| log(ALT) | 0.701 | 0.042 to 1.360 | .037 | .084 | .014 |
| XL probe | 0.796 | −0.094 to 1.685 | .079 | .125 | .010 |
| VCTE IQR/median, per percentage point | −0.006 | −0.048 to 0.037 | .788 | .803 | .000 |
| 2D-SWE IQR/median, per percentage point | 0.021 | −0.041 to 0.082 | .512 | .584 | .001 |
| LOGIQ E10 device | −1.092 | −1.916 to −0.267 | .010 | .030 | .021 |
| Mean LSM, per kPa | 0.998 | 0.925 to 1.071 | < .001 | < .001 | .698 |

**Panel B. Multivariable model**

| Predictor | Adjusted β, kPa | 95% CI | *P* value | BH q value |
|---|---|---|---|---|
| Age, per year | −0.034 | −0.056 to −0.013 | .002 | .009 |
| Female sex | 0.367 | −0.093 to 0.827 | .118 | .179 |
| Waist circumference, per cm | −0.009 | −0.028 to 0.010 | .336 | .394 |
| CAP, per dB/m | 0.004 | −0.004 to 0.012 | .319 | .385 |
| log(ALT) | −0.585 | −1.141 to −0.029 | .039 | .084 |
| XL probe | −0.298 | −0.836 to 0.240 | .277 | .344 |
| LOGIQ E10 device | 0.280 | −0.173 to 0.733 | .225 | .288 |
| Mean LSM, per kPa | 1.062 | 0.937 to 1.186 | < .001 | < .001 |

Signed discrepancy was defined as VCTE LSM − 2D-SWE LSM. Positive coefficients indicate increasingly higher VCTE values relative to 2D-SWE; negative coefficients indicate a smaller positive difference or relatively higher 2D-SWE values. Panel A reports separate univariable linear models. Panel B includes all listed predictors simultaneously; BMI was not included with waist circumference to avoid collinearity. Mean LSM is mathematically related to the signed difference and its coefficient should therefore be interpreted descriptively, not causally. BH q values are Benjamini–Hochberg false-discovery-rate-adjusted values from the prespecified pool of 41 exploratory tests. Reference categories were male sex, M probe, and LOGIQ E9. 2D-SWE, two-dimensional shear-wave elastography; ALT, alanine aminotransferase; BH, Benjamini–Hochberg; BMI, body mass index; CAP, controlled attenuation parameter; CI, confidence interval; IQR, interquartile range; LSM, liver stiffness measurement; $R^2$, coefficient of determination; VCTE, vibration-controlled transient elastography.
