## Supplementary Figures for "Switching Between VCTE and 2D-SWE Reclassifies Fibrosis Risk in Metabolic Dysfunction-Associated Steatotic Liver Disease"

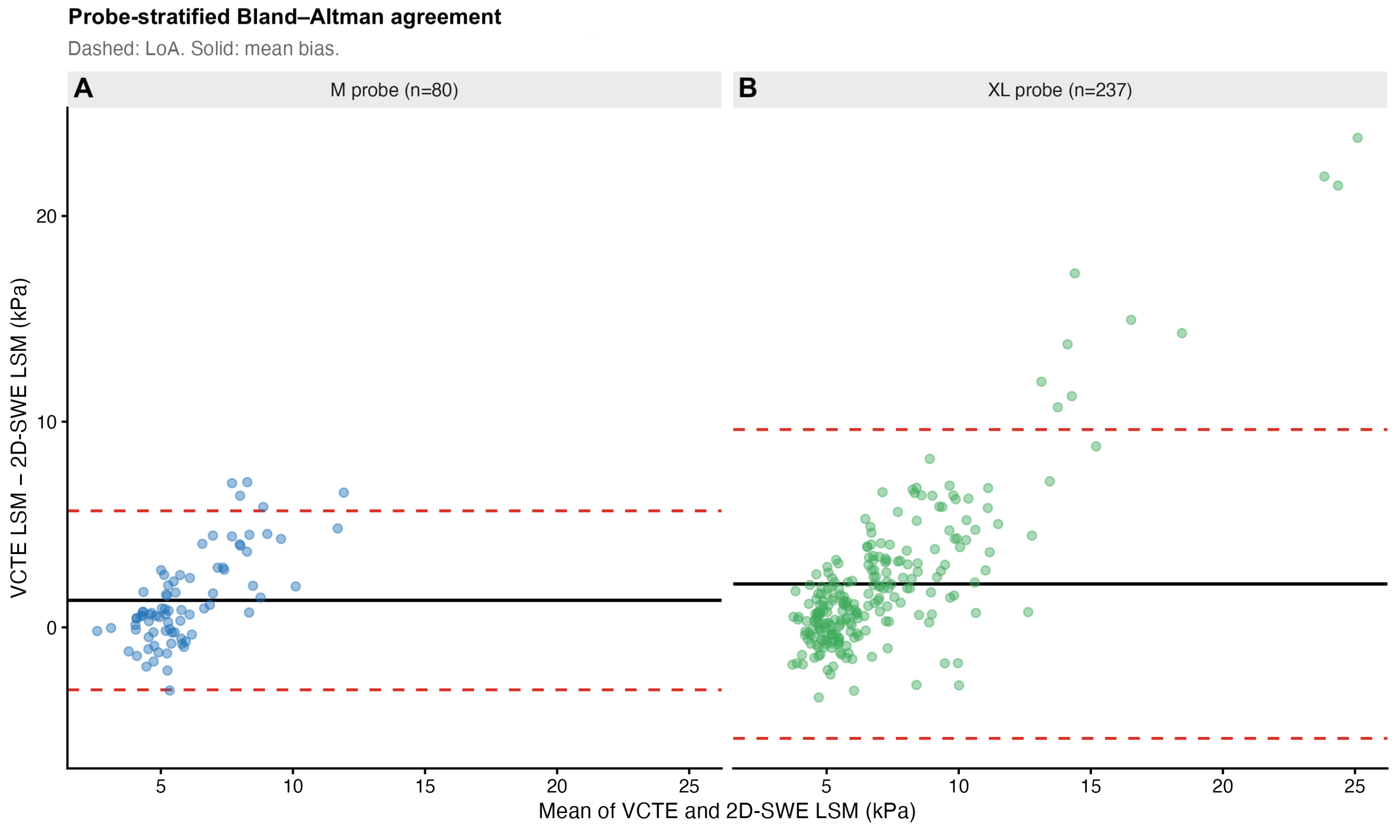


**Supplementary Figure S1.** Bland–Altman agreement stratified by VCTE probe type. Bland–Altman plots show the paired VCTE–2D-SWE liver-stiffness difference against the mean of the two measurements for examinations performed using the VCTE M probe (Panel A, n = 80) and XL probe (Panel B, n = 237). Solid black lines indicate the mean inter-method bias, and dashed red lines indicate the 95% limits of agreement. For M-probe examinations, the mean bias was 1.31 kPa, with limits of agreement from −3.04 to 5.66 kPa. For XL-probe examinations, the mean bias was 2.11 kPa, with wider limits of agreement from −5.40 to 9.62 kPa. Positive differences indicate higher liver-stiffness measurements by VCTE than by 2D-SWE. Probe selection followed clinical criteria and was associated with patient body habitus; therefore, the probe-stratified differences are descriptive and should not be interpreted as an independent causal effect of probe type. 2D-SWE, two-dimensional shear-wave elastography; VCTE, vibration-controlled transient elastography.

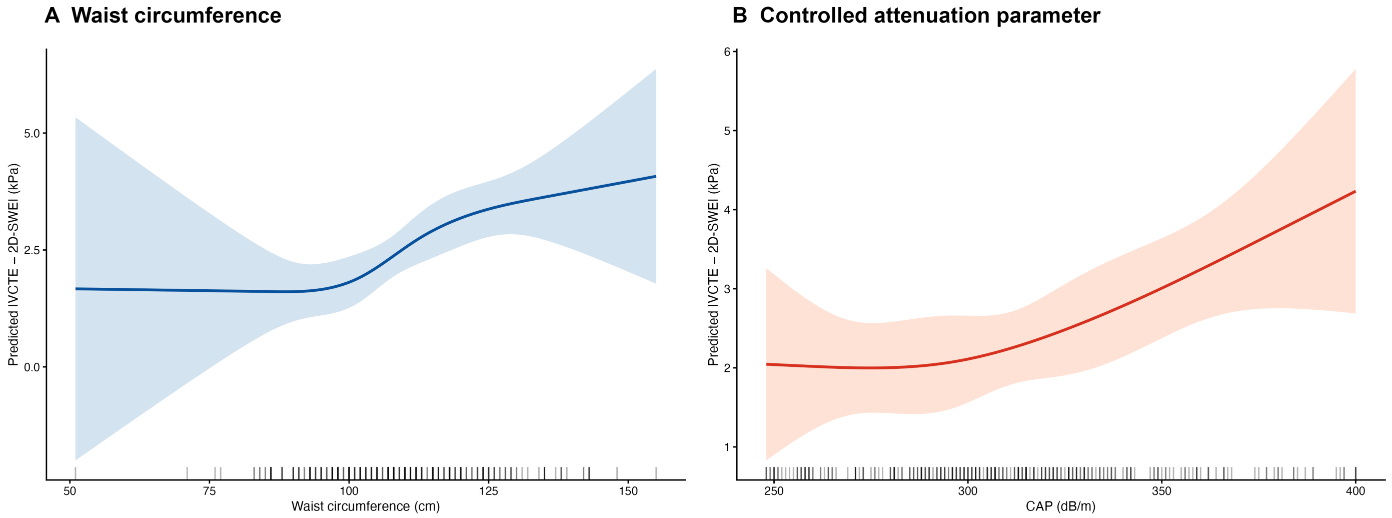


**Supplementary Figure S2.** Exploratory associations of waist circumference and controlled attenuation parameter with absolute inter-method discrepancy. Restricted cubic spline models show the predicted absolute VCTE–2D-SWE difference according to waist circumference (Panel A) and controlled attenuation parameter (Panel B). Solid lines indicate model-predicted absolute differences, shaded areas indicate 95% confidence intervals, and rug marks show the distributions of observed predictor values. Both models had low explanatory power ($R^2 = 0.049$ for waist circumference and $R^2 = 0.029$ for controlled attenuation parameter), and confidence intervals widened substantially at distribution extremes where observations were sparse. The panels retain independent y-axis ranges and should not be compared on the basis of apparent curve steepness. Because the spline models were fitted using unconstrained linear predictions, confidence limits may extend below zero despite the non-negative nature of an absolute difference. These analyses were exploratory, and controlled attenuation parameter was derived from the VCTE platform. CAP, controlled attenuation parameter; 2D-SWE, two-dimensional shear-wave elastography; VCTE, vibration-controlled transient elastography.

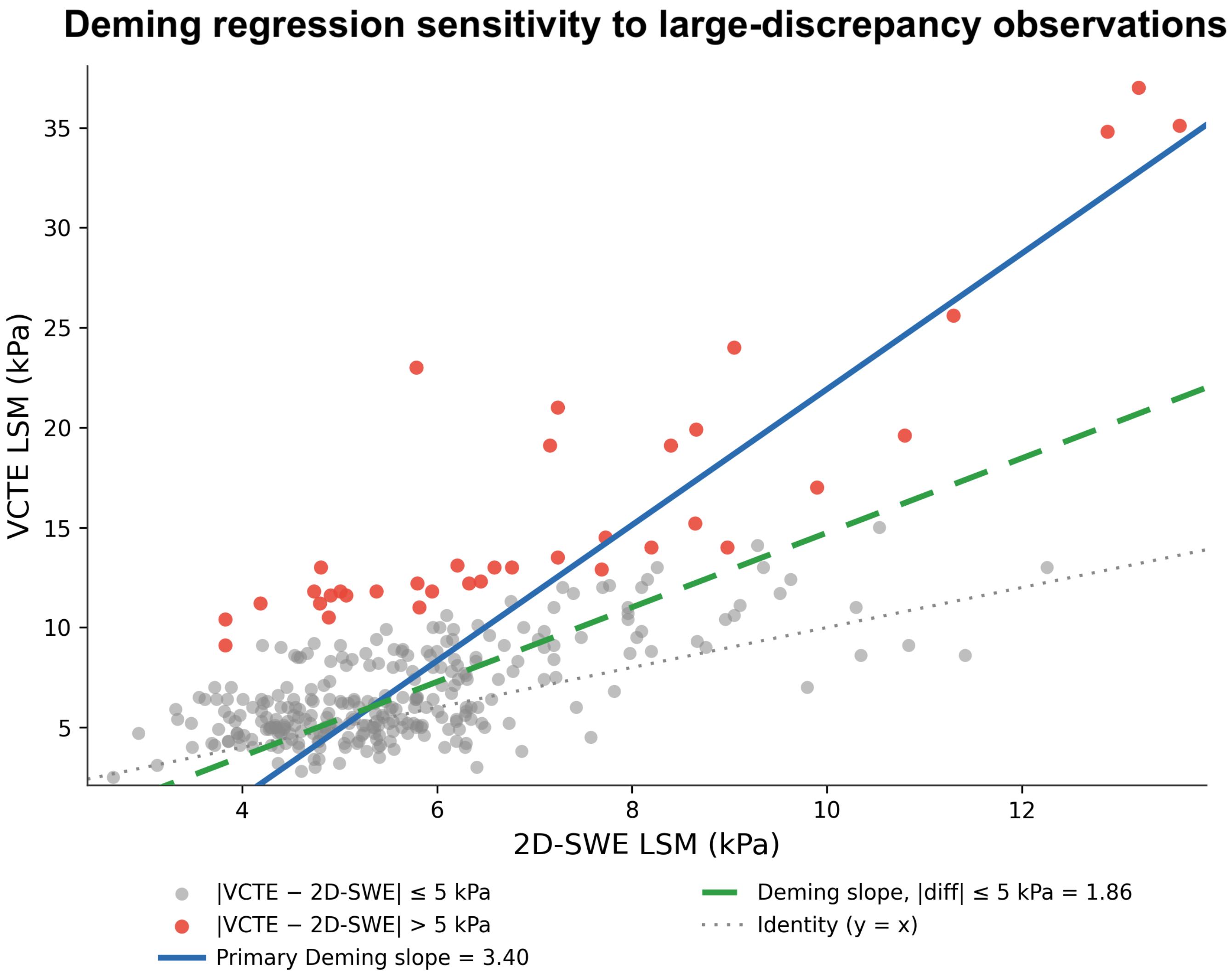


**Supplementary Figure S3.** Sensitivity of Deming regression to large-discrepancy observations. The scatterplot shows paired VCTE and 2D-SWE liver-stiffness measurements. The solid blue line represents the primary Deming regression slope of 3.40, and the dashed green line represents the Deming regression slope of 1.86 after restricting the analysis to observations with an absolute VCTE–2D-SWE difference ≤5 kPa. The dotted grey line represents identity. Grey points met the ≤5-kPa restriction, whereas red points had an absolute inter-method difference >5 kPa and were excluded only from the restricted sensitivity analysis. Although exclusion of large-discrepancy observations substantially reduced the slope estimate, the restricted slope remained above unity, supporting persistent scale non-identity while demonstrating that the magnitude of the primary Deming slope was leverage-sensitive. 2D-SWE, two-dimensional shear-wave elastography; VCTE, vibration-controlled transient elastography.

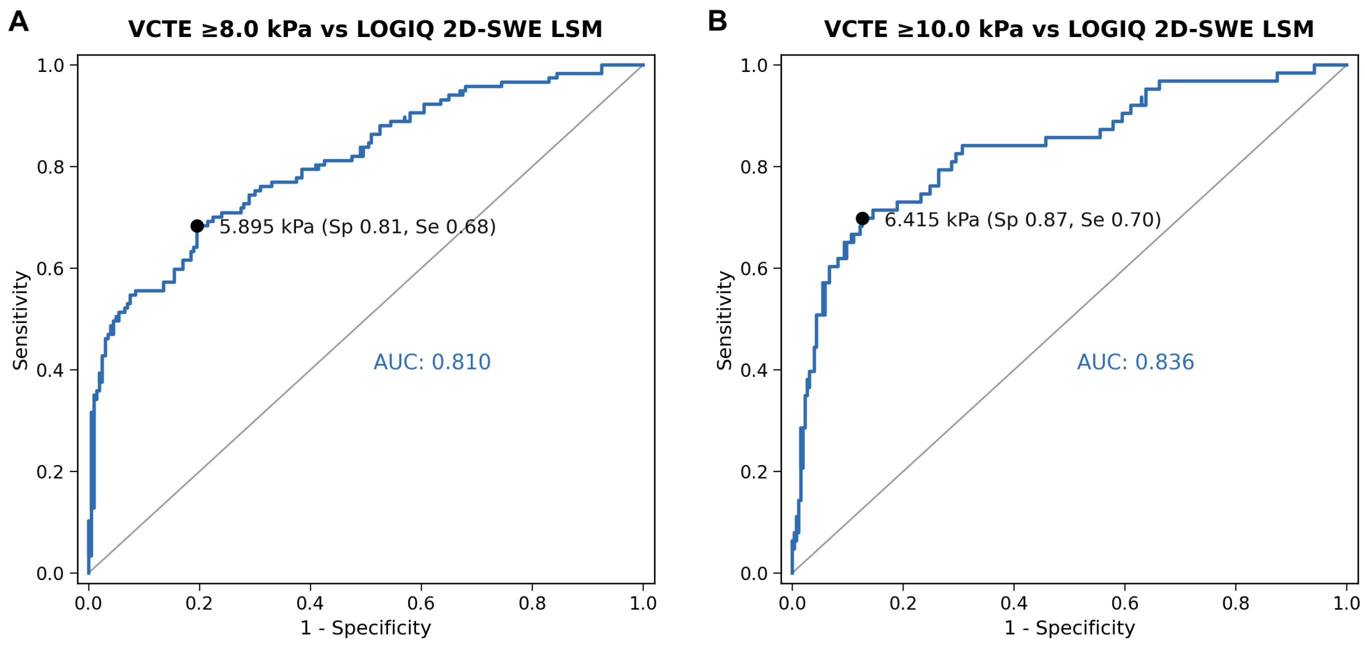


**Supplementary Figure S4.** Receiver operating characteristic curves for VCTE-referenced operational threshold recalibration. Curves show the ability of continuous GE LOGIQ 2D-SWE liver-stiffness measurements to discriminate patient classifications defined by the prespecified VCTE decision thresholds. Panel A shows the lower VCTE threshold of ≥8.0 kPa. The area under the receiver operating characteristic curve was 0.810 (95% CI, 0.76–0.86), and the apparent Youden-derived 2D-SWE threshold was 5.895 kPa, with sensitivity 0.68 and specificity 0.81. Panel B shows the upper VCTE threshold of ≥10.0 kPa. The area under the curve was 0.836 (95% CI, 0.77–0.90), and the apparent Youden-derived 2D-SWE threshold was 6.415 kPa, with sensitivity 0.70 and specificity 0.87. Black points identify the apparent Youden-derived thresholds, and grey diagonal lines indicate chance discrimination. VCTE classification was used as an operational reference for method harmonisation, not as a histological or diagnostic reference standard; consequently, the curves should not be interpreted as estimates of diagnostic accuracy for liver fibrosis. Bootstrap confidence intervals and internal-validation results for the recalibrated thresholds are reported in Supplementary Table S11. 2D-SWE, two-dimensional shear-wave elastography; VCTE, vibration-controlled transient elastography.

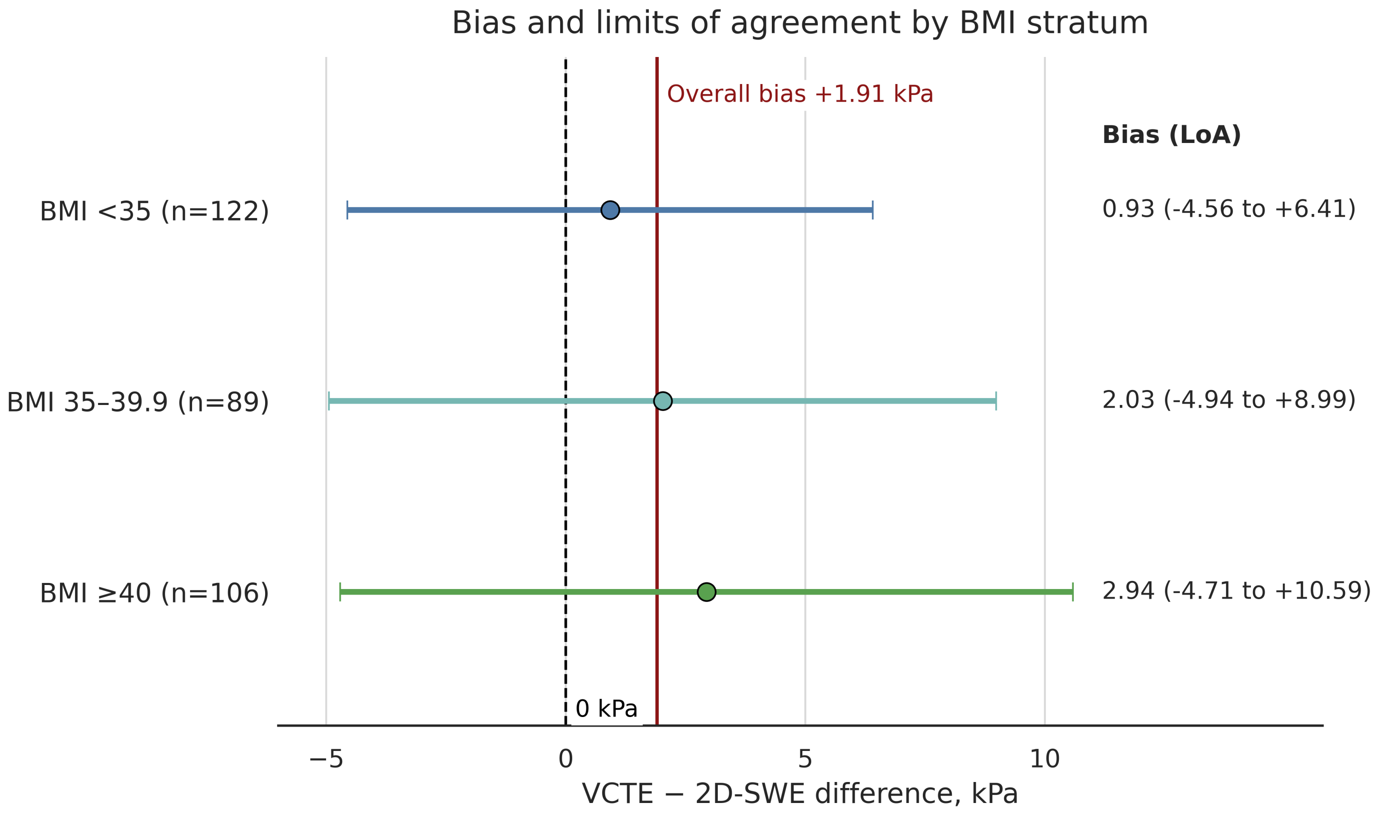


**Supplementary Figure S5.** Bias and limits of agreement by BMI stratum. Points indicate the mean paired VCTE − 2D-SWE difference, and horizontal bars indicate the 95% limits of agreement within each BMI stratum. Positive values indicate higher liver-stiffness measurements by VCTE than by 2D-SWE. Mean bias increased across BMI strata from 0.93 kPa in patients with BMI <35 kg/m² to 2.94 kPa in patients with BMI ≥40 kg/m², with progressive widening of the upper limit of agreement. The dashed vertical line denotes zero difference, and the red vertical line denotes the overall mean bias of 1.91 kPa. BMI, body mass index; 2D-SWE, two-dimensional shear-wave elastography; VCTE, vibration-controlled transient elastography.
